# Genetic QT Score and Sleep Apnea as Predictors of Sudden Cardiac Death in the UK Biobank

**DOI:** 10.1101/2023.11.07.23298237

**Authors:** Amit Arora, Wojciech Zareba, Raymond L. Woosley, Yann C. Klimentidis, Imran Y. Patel, Stuart F. Quan, Christopher Wendel, Fadi Shamoun, Stefano Guerra, Sairam Parthasarathy, Salma I. Patel

## Abstract

**Introduction:** The goal of this study was to evaluate the association between a polygenic risk score (PRS) for QT prolongation (QTc-PRS), QTc intervals and mortality in patients enrolled in the UK Biobank with and without sleep apnea.

**Methods:** The QTc-PRS was calculated using allele copy number and previously reported effect estimates for each single nuclear polymorphism SNP. Competing-risk regression models adjusting for age, sex, BMI, QT prolonging medication, race, and comorbid cardiovascular conditions were used for sudden cardiac death (SCD) analyses.

**Results:** 500,584 participants were evaluated (56.5 ±8 years, 54% women, 1.4% diagnosed with sleep apnea). A higher QTc-PRS was independently associated with the increased QTc interval duration (p<0.0001). The mean QTc for the top QTc-PRS quintile was 15 msec longer than the bottom quintile (p<0.001). Sleep apnea was found to be an effect modifier in the relationship between QTc-PRS and SCD. The adjusted HR per 5-unit change in QTc-PRS for SCD was 1.64 (95% CI 1.16 – 2.31, p=0.005) among those with sleep apnea and 1.04 (95% CI 0.95 – 1.14, p=0.44) among those without sleep apnea (p for interaction =0.01). Black participants with sleep apnea had significantly elevated adjusted risk of SCD compared to White participants (HR=9.6, 95% CI 1.24 - 74, p=0.03).

**Conclusion:** In the UK Biobank population, the QTc-PRS was associated with SCD among participants with sleep apnea but not among those without sleep apnea, indicating that sleep apnea is a significant modifier of the genetic risk. Black participants with sleep apnea had a particularly high risk of SCD.

## INTRODUCTION

Obstructive sleep apnea is a prevalent medical condition characterized by repetitive upper airway collapse.^1^ Patients with obstructive sleep apnea have prolonged QTc intervals during the daytime which further increases during sleep when apnea/hypopnea events occur.^1–5^ Prolongation of the corrected QT interval (QTc) in electrocardiograms is associated with increased risk for ventricular arrhythmias, sudden cardiac death (SCD), and all-cause mortality.^6–20^ Up to 45% of the variation in the QTc interval has been attributed to genetic factors.^21–23^ However, the impact of these genetic factors has yet to be explored in patients with obstructive sleep apnea.

Genome-wide association studies (GWASs) have identified common genetic variants that associate with differences in QTc intervals.^22,24–26^ The QTGEN and QTSCD consortia established 15 independent single nucleotide polymorphisms (SNPs) with a genome-wide significant association with QTc duration.^22,26^ Noseworthy et al. incorporated these SNPs into an overall polygenic risk score (QTc-PRS) by using the allele copy number and previously reported effect estimates for each SNP, and found the QTc-PRS score to be a continuous independent predictor of the QTc interval (P<10^-107^), with patients in the top quintile having a 15 ms higher QTc interval compared to those in the bottom quintile.^25^ The association of the QT score with SCD risk was noted to be U-shaped, with the risk of SCD higher in the top QT score quintile compared with the middle quintile.^25^

Currently, approximately 68 independent SNPs at 35 loci have been identified and are thought to explain 7.6-9.9% of the variance in the QTc interval.^24^ The QTc-PRS created with SNPs at these 35 loci also explain a significant proportion of the variability in drug-induced QT prolongation, as well as drug-induced *torsade de pointes* leading to sudden cardiac death.^27^ Patients with sleep apnea may particularly be affected by increased QTc-PRS given a longer QTc interval in their resting daytime electrocardiograms^2^ and further prolongation in sleep during apnea/hypopnea events.^3,4^ However, the relationship of a QTc-PRS with SCD has not been examined among this group of at-risk individuals.

The goal of this study was to test whether an increased QTc-PRS is associated with an increased risk of SCD in participants enrolled in the UK Biobank and if this relationship is affected by the presence of sleep apnea.

## METHODS

### Population and Variables

The study was approved by the University of Arizona Institutional Review Board and additionally a data use agreement was obtained from the UK Biobank. The UK Biobank has information for approximately 500,000 volunteer participants with ages 40-69 years recruited from cities across the United Kingdom between 2006-2010 with long-term follow-up data.^28^ The UK BiLEVE array and UK Biobank Axiom Arrays are well validated and were used to analyze the genetic data for these subjects. The UK BiLEVE array was run on the first approximately 50,000 samples genotyped for UK Biobank.^21^ The UK BiLEVE and UK Biobank Axiom Arrays are similar with over 95% common content.^29^ There are 820,967 SNP and indel markers on these arrays.^29^ Death information is kept up to date in the UK Biobank through linkage with the national death registries which provide date and cause of death according to the International Classification of Diseases-10 (ICD-10) if the death occurred in the UK.^29^

Variables collected for analysis from this dataset included age, sex, body mass index (BMI), self-reported race, diagnosed sleep apnea, QT prolonging medications, comorbid cardiovascular conditions, SNP data, date of intake assessment, and date and cause of death. In addition, QTc duration was available for roughly 5% of the sample with QT-PRS. QT Prolonging medications with known risk of Torsades de Pointes (TdP) were consistent with the Arizona CredibleMeds QT drug list. ^1,30^ Comorbid cardiovascular conditions was a dichotomous variable indicating history of any of the following: myocardial infarction, angina, stroke, or high blood pressure. QTc duration calculated using the Bazett’s heart rate correction formula was also collected for all participants that had undergone 12-lead electrocardiograms at rest, which was a small subset of the entire sample. Cause of death was considered SCD for cases in which the primary or secondary cause of death was listed as sudden cardiac death (ICD10 I46.1) or cardiac arrest (ICD10 I46.9). Race was self-reported from choices White, Black, Asian, Chinese, Mixed, Other, and Not Reported. Due to relatively small numbers of Chinese patients (n=1,503), we combined them with Asian.

### Statistical analysis

All analysis included adjustment for age, sex, BMI, race, QT prolonging medication and comorbid cardiovascular conditions. To explore whether the QTc-PRS was associated with SCD and all-cause mortality in the UK Biobank sample, we used the method established by Noseworthy et al.^25^ to conduct these analyses. The QTc-PRS for all patients was calculated using the allele copy number and the previously established effect estimates^24^ for the 68 SNPs using the following formula: QTc-PRS = [(SNP1 allele copy number) × (SNP1 effect estimate in predicted msec)] + [(SNP2 allele copy number) × (SNP2 effect estimate in predicted msec)] +…through all 68 SNPs.^25^ SNP genotypes that were missing were calculated to have allele copy number equal to two times the coded allele frequency in the total sample.^25,31^ The genotype score is expressed in “predicted msec” of expected change in the QT interval.^25^

We calculated the correlation between QTc duration and QTc-PRS using Pearson’s r-square, and determined the proportion of variation in QTc explained by QTc-PRS using the squared semipartial correlation coefficient after removing effects of age, sex, BMI, race, QT prolonging medication and comorbid cardiovascular conditions. We used a nonparametric test for trend across ordered groups developed by Cuzick (1985) to evaluate the trend of mean QTc duration across QTc-PRS quintiles.^32^ Because QTc duration was available only for the 5% subset of the UKB patients who had undergone 12-lead electrocardiograms at rest, we did not treat absence of QTc duration as missing requiring imputation. For survival analysis, the exposure of interest was the continuous QTc-PRS. For significant associations we evaluated the point estimates for quintiles of QTc-PRS to judge whether the association appeared to change linearly with QTc-PRS. Missing data was minimal for covariates in the survival analysis regression. BMI was missing for 0.40%, comorbid cardiovascular conditions for 0.36%, and race for 0.43%. However, we kept the race category of “Not reported” in survival models, so it was effectively not missing in survival models and the risk for that group was calculated. The total missing either BMI or comorbid cardiovascular conditions was less than 0.75%, which we believed did not warrant imputation approaches for missing data sensitivity analysis.

The time interval of observation was defined as the duration from date of attending assessment center to death date, date lost to follow up or when the data were last downloaded from the UK Biobank (6/2020). We performed competing-risks regression as developed by Fine and Gray (1999),^34^ which is a maximum likelihood alternative to Cox proportional hazards regression that accounts for competing risks from causes of death other than the failure outcome. Competing-risks regression (Stata command “stcrreg”) on the specified cause of death categories were performed with all deaths other than the specified cause designated as competing-risk events adjusted for potential confounders age, sex, BMI, race, QT prolonging medication and comorbid cardiovascular conditions. Results are presented as the hazard ratio (HR), which is interpreted as a relative risk based on time to death per 5-unit change in QTc-PRS, along with standard error of HR, p-value of HR, and 95% confidence interval of HR. For context of a 5-unit change in QTc-PRS, the standard deviation of the QTc-PRS is 7.4, the gap between 10^th^ and 50^th^ percentile is 9.4, and the gap between 50^th^ and 90^th^ percentile is 9.6. The Competing-risk regression models were performed with continuous QTc-PRS for hypothesis testing, and with quintiles of QTc-PRS for assessment of linear relationship with risk of death. For single SNP to QTc interval analyses, the threshold of significance was set at p<0.0007 based on Bonferroni correction (0.05/68), and for other analyses significance was defined as p<0.05.^25^

A sensitivity analyses was done by taking into consideration additional causes of death similar to Noseworthy et al ^25^ including probable SCD (primary or secondary cause of death was any other conduction disorder, possible SCD (primary or secondary cause of death was cardiovascular but not arrhythmia related) and “other” including those that could have reasonably been attributed to an arrhythmia^25^ due to the potential uncertainty associated with some of these situations, as no manual adjudication was performed per participant.

## RESULTS

### Demographics

Demographic and clinical characteristics of all the participants in the UK Biobank based on mortality status are described in Table 1. There were 223 SCD and 19,589 deaths from all causes. Age, male sex, BMI, presence of sleep apnea and comorbid cardiovascular conditions had significantly higher mean or proportion in both SCD vs. No SCD and all-cause mortality vs. alive (p<0.01). For race, the proportion White was significantly higher for SCD vs. No SCD, but lower for all-cause mortality vs. alive (p<0.01). QT prolonging medication had a significantly higher frequency in all-cause mortality vs. alive (p<0.01).

**Table 1.**
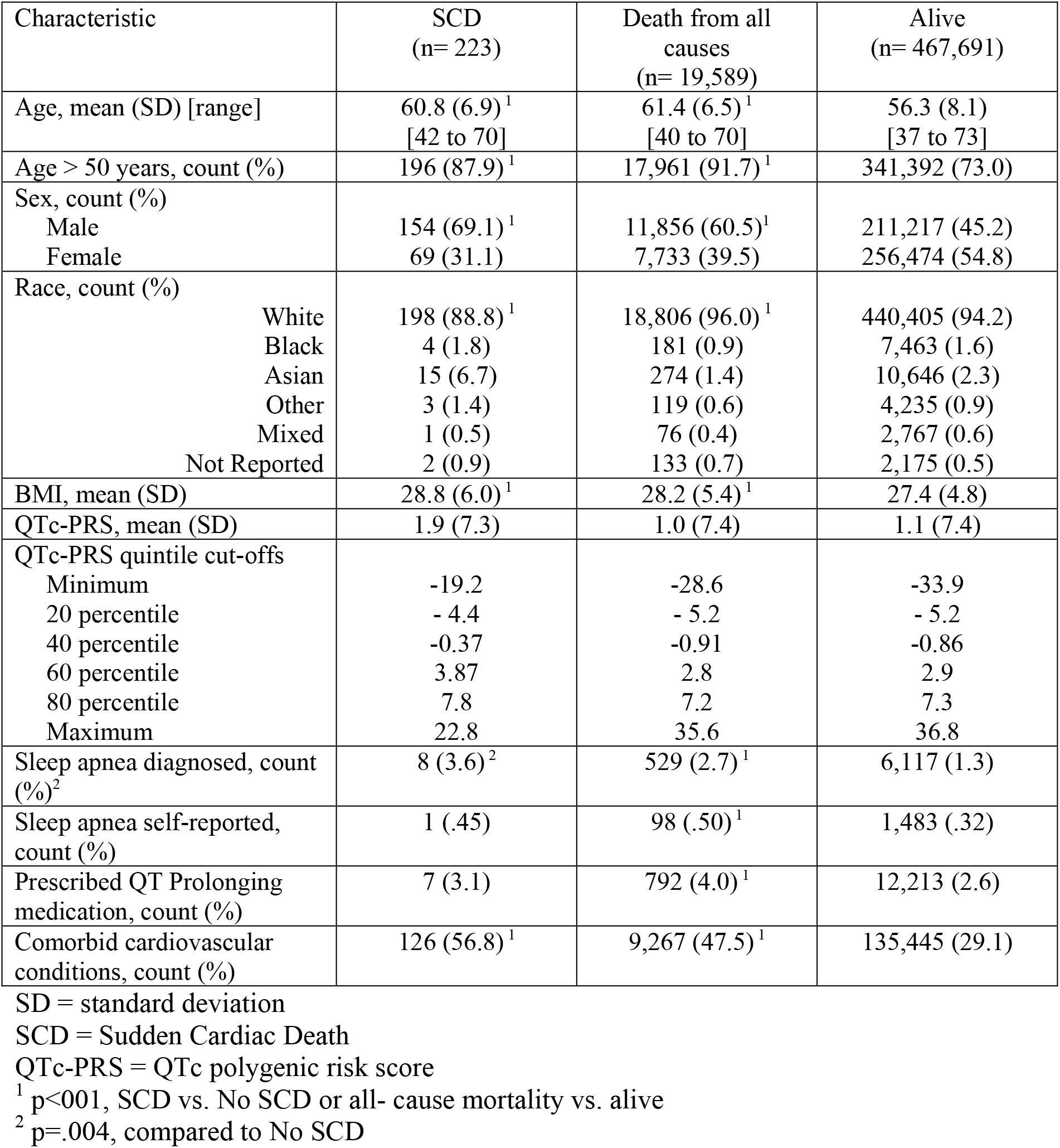
Participant Demographics and Mortality in the UK Biobank.

### QTc duration and QTc-PRS

QTc duration was available for 24,924 participants. The individual contribution of the SNPs to QTc duration are demonstrated in Supplementary Table 1. The following SNPs were associated (p<0.0007) with QTc duration when considered individually: rs12079745, rs3857067, rs1961102 and rs1052536. The mean QTc for the top QTc-PRS quintile was 15 msec longer than the bottom quintile (p<0.001). QTc duration is plotted against the QTc-PRS quintiles in Figure 1 and shows a linear relationship with QTc-PRS. When analyzed separately by sex, the Pearson correlation for men is r=0.18, p<0.0001 and for women is r=0.20, p<0.0001. Means for both sexes climb steadily across quintiles (with means for women roughly 10 ms higher than in men in respective quintiles) and have a significant trend (p<0.001 in men and women, respectively). The QTc-PRS explained 3.7% (p<.0001) of the variation in QTc after adjustment for age, sex, BMI, race, QT prolonging medication and comorbid cardiovascular conditions. The Pearson correlation between QTc duration and QTc-PRS for both sexes combined was significant in those with and without sleep apnea (p<0.0001). QTc duration is plotted against the QTc-PRS quintiles in Figure 2a and 2b for participants with and without sleep apnea, respectively.

**Figure 1:**
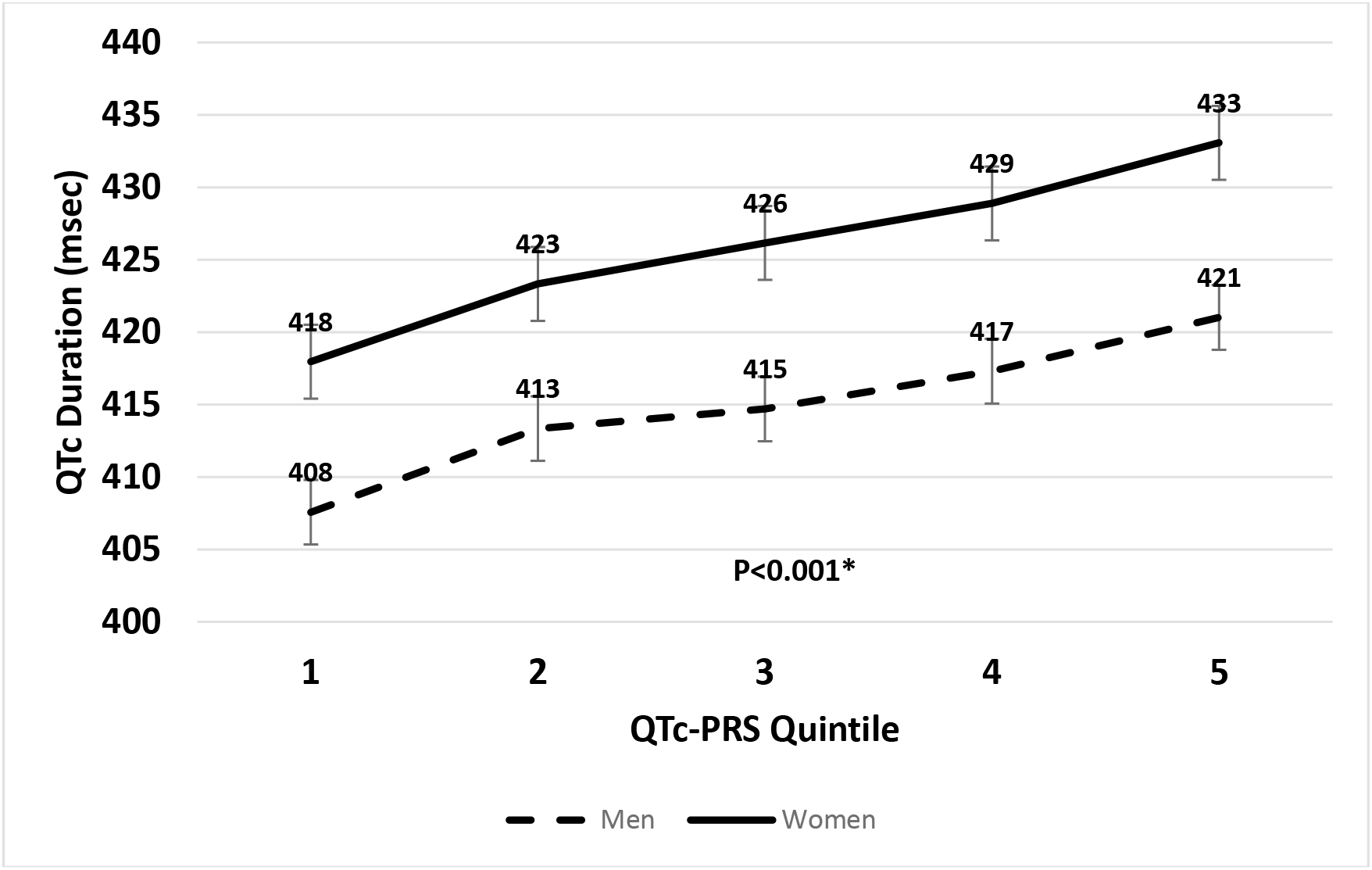
QTc duration correlation with QTc PRS quintiles in men and women.

**Figure 2:**
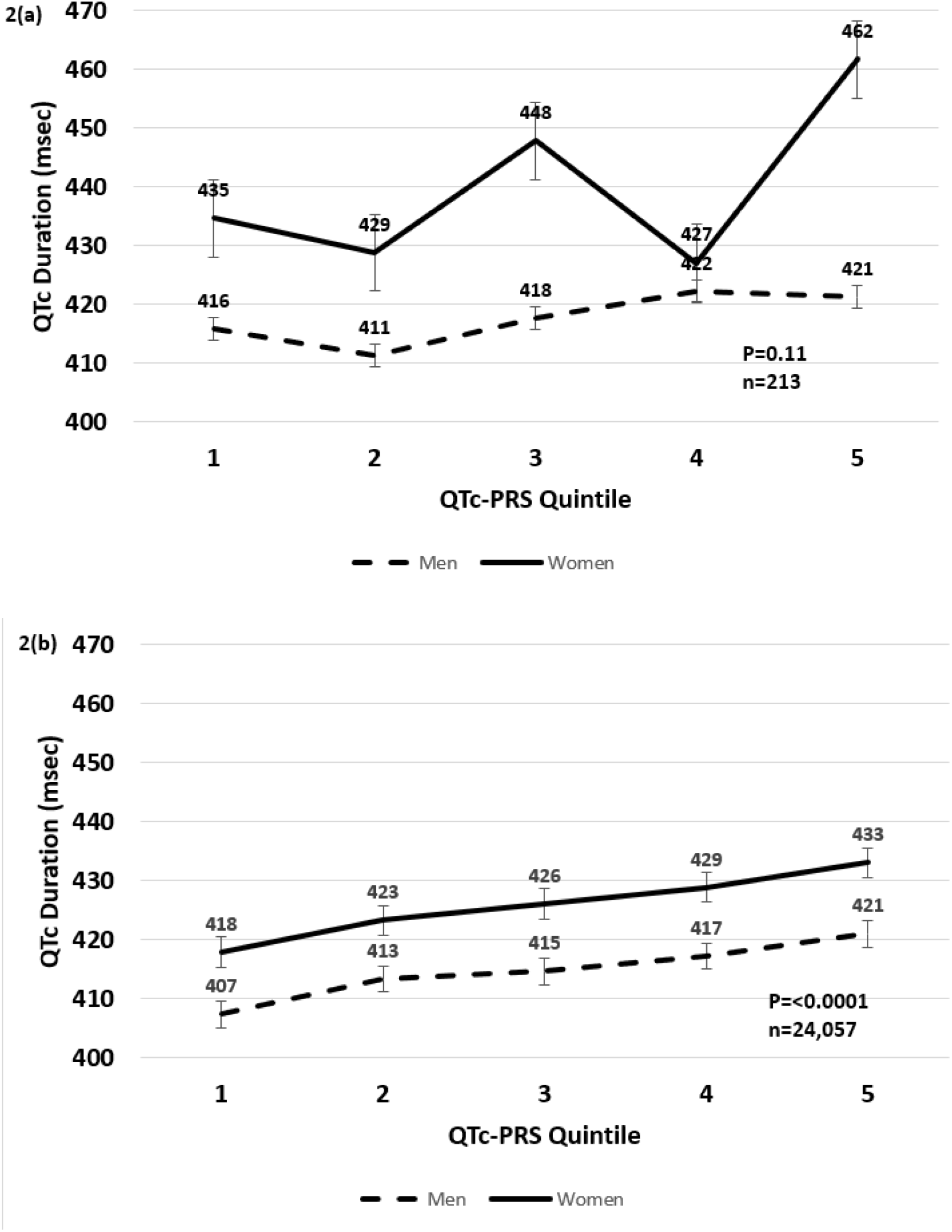
QTc duration correlation with QTc PRS quintiles in men and women (a) with and (b) without sleep apnea.

### QTc-PRS and Mortality

The individual associations with SCD for of the SNPs contributing to the QTc-PRS score are demonstrated in Supplementary Table 1. The following SNPs were nominally associated with SCD risk when considered individually: rs3934467 and rs3857067. The adjusted risk of death from SCD did not monotonically rise over quintiles of QTc-PRS, referenced to quintile 1 (0.91, p=0.68; 1.15, p=0.52; 1.22, p=0.37; 1.13, p=0.57; respectively). This pattern suggests that there is no change between quintiles 1 and 2, increases for 3 and again for 4, and plateau or decrease between 4 and 5. Table 2 shows that QTc-PRS was not significantly associated with SCD or all-cause death in unadjusted and adjusted models.

**Table 2.** Risk of death associated with QTc-PRS.

|  | **Unadjusted** |  |  |  | **Adjusted<sup>2</sup>** |  |  |  |
| --- | --- | --- | --- | --- | --- | --- | --- | --- |
| **Cause of death** | **HR<sup>1</sup>** | **SE** | **p-value** | **95% CI** | **HR<sup>1</sup>** | **SE** | **p-value** | **95% CI** |
| SCD | 1.08 | 0.05 | 0.10 | 0.99 - 1.17 | 1.05 | 0.05 | 0.29 | 0.96 - 1.15 |
| All causes | 1.00 | 0.005 | 0.31 | 0.99 - 1.00 | 1.00 | 0.005 | 0.68 | 0.99 - 1.01 |

HR = hazard ratio
SE = standard error
CI = confidence interval
QTc-PRS = QTc polygenic risk score
SCD = Sudden Cardiac Death
<sup>1</sup> HR per 5-unit change in continuous QTc-PRS
<sup>2</sup> Adjusted for age, sex, race, BMI, QT Prolonging medication, and comorbid cardiovascular conditions

### QTc-PRS, Mortality and Sleep Apnea

Table 3 shows the risk of SCD and all-cause death stratified by physician-diagnosed sleep apnea. There were 6,646 (1.36%) patients with physician-diagnosed sleep apnea. Of those who self-reported sleep apnea (1,581 or 0.32%), there was a subset without physician-diagnosed sleep apnea, but inclusion of the self-reported patients in the analysis did not appreciably change the model results.

**Table 3.**
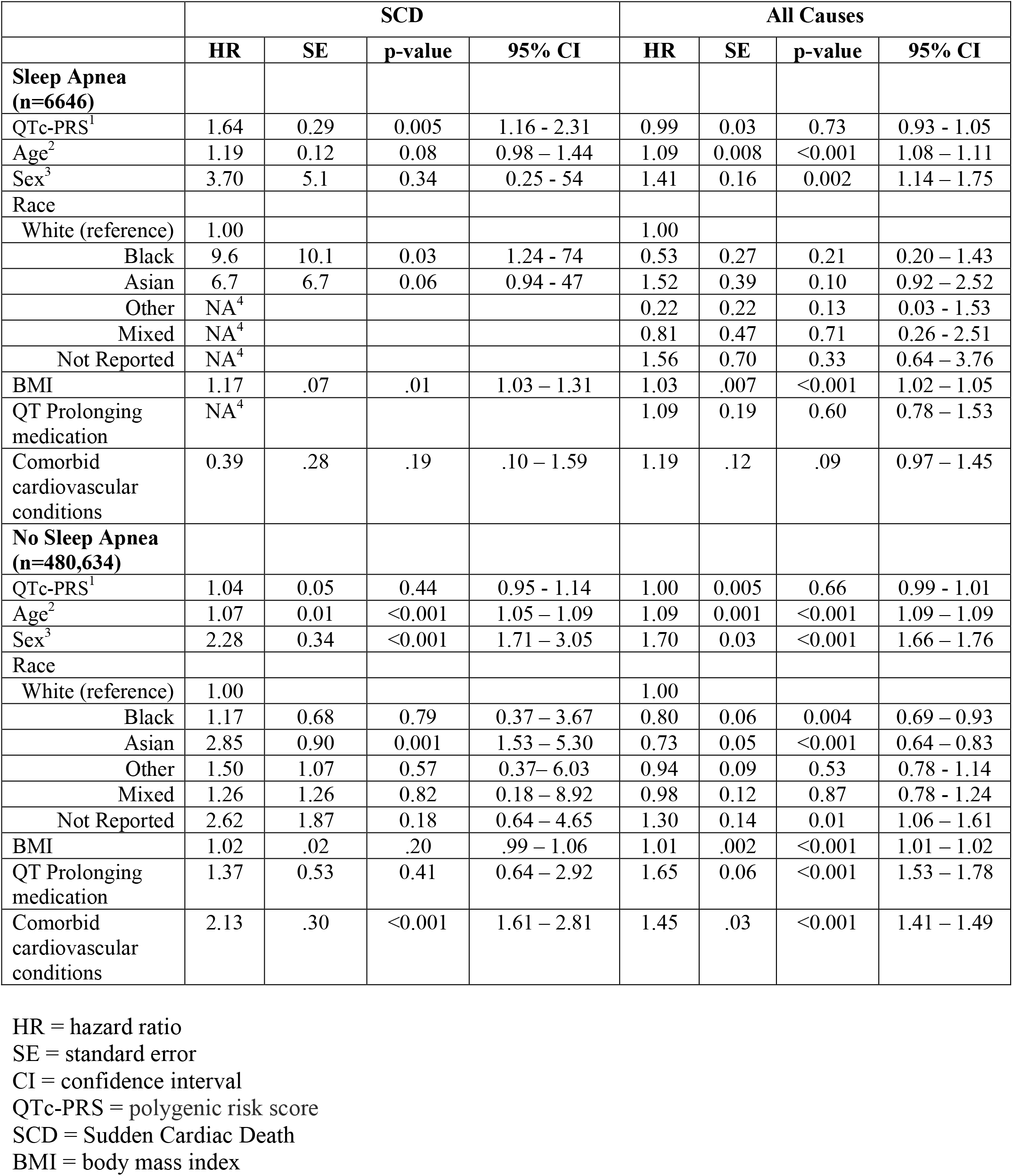
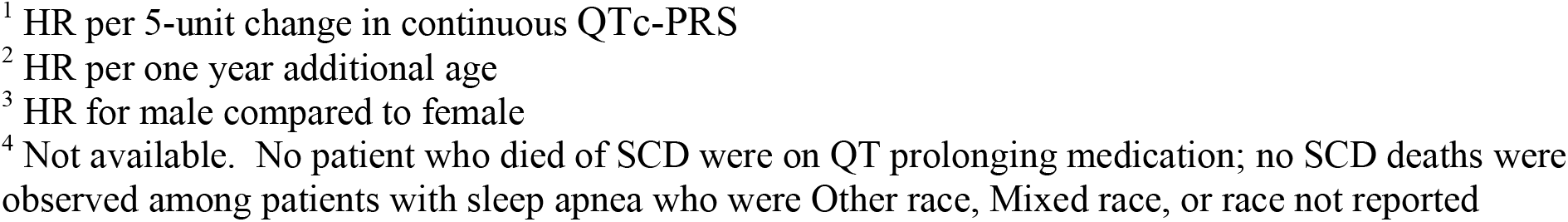
Hazard Ratios for SCD and all-cause death in multivariate models stratified by the presence of sleep apnea.

|  | SCD |  |  |  | All Causes |  |  |  |
| --- | --- | --- | --- | --- | --- | --- | --- | --- |
|  | HR | SE | p-value | 95% CI | HR | SE | p-value | 95% CI |
| **Sleep Apnea**<br>**(n=6646)** |  |  |  |  |  |  |  |  |
| QTc-PRS<sup>1</sup> | 1.64 | 0.29 | 0.005 | 1.16 - 2.31 | 0.99 | 0.03 | 0.73 | 0.93 - 1.05 |
| Age<sup>2</sup> | 1.19 | 0.12 | 0.08 | 0.98 – 1.44 | 1.09 | 0.008 | <0.001 | 1.08 – 1.11 |
| Sex<sup>3</sup> | 3.70 | 5.1 | 0.34 | 0.25 - 54 | 1.41 | 0.16 | 0.002 | 1.14 – 1.75 |
| Race |  |  |  |  |  |  |  |  |
| White (reference) | 1.00 |  |  |  | 1.00 |  |  |  |
| Black | 9.6 | 10.1 | 0.03 | 1.24 - 74 | 0.53 | 0.27 | 0.21 | 0.20 – 1.43 |
| Asian | 6.7 | 6.7 | 0.06 | 0.94 - 47 | 1.52 | 0.39 | 0.10 | 0.92 – 2.52 |
| Other | NA<sup>4</sup> |  |  |  | 0.22 | 0.22 | 0.13 | 0.03 - 1.53 |
| Mixed | NA<sup>4</sup> |  |  |  | 0.81 | 0.47 | 0.71 | 0.26 - 2.51 |
| Not Reported | NA<sup>4</sup> |  |  |  | 1.56 | 0.70 | 0.33 | 0.64 – 3.76 |
| BMI | 1.17 | .07 | .01 | 1.03 – 1.31 | 1.03 | .007 | <0.001 | 1.02 – 1.05 |
| QT Prolonging medication | NA<sup>4</sup> |  |  |  | 1.09 | 0.19 | 0.60 | 0.78 – 1.53 |
| Comorbid cardiovascular conditions | 0.39 | .28 | .19 | .10 – 1.59 | 1.19 | .12 | .09 | 0.97 – 1.45 |
| **No Sleep Apnea**<br>**(n=480,634)** |  |  |  |  |  |  |  |  |
| QTc-PRS<sup>1</sup> | 1.04 | 0.05 | 0.44 | 0.95 - 1.14 | 1.00 | 0.005 | 0.66 | 0.99 - 1.01 |
| Age<sup>2</sup> | 1.07 | 0.01 | <0.001 | 1.05 – 1.09 | 1.09 | 0.001 | <0.001 | 1.09 – 1.09 |
| Sex<sup>3</sup> | 2.28 | 0.34 | <0.001 | 1.71 – 3.05 | 1.70 | 0.03 | <0.001 | 1.66 – 1.76 |
| Race |  |  |  |  |  |  |  |  |
| White (reference) | 1.00 |  |  |  | 1.00 |  |  |  |
| Black | 1.17 | 0.68 | 0.79 | 0.37 – 3.67 | 0.80 | 0.06 | 0.004 | 0.69 – 0.93 |
| Asian | 2.85 | 0.90 | 0.001 | 1.53 – 5.30 | 0.73 | 0.05 | <0.001 | 0.64 – 0.83 |
| Other | 1.50 | 1.07 | 0.57 | 0.37– 6.03 | 0.94 | 0.09 | 0.53 | 0.78 - 1.14 |
| Mixed | 1.26 | 1.26 | 0.82 | 0.18 – 8.92 | 0.98 | 0.12 | 0.87 | 0.78 - 1.24 |
| Not Reported | 2.62 | 1.87 | 0.18 | 0.64 – 4.65 | 1.30 | 0.14 | 0.01 | 1.06 – 1.61 |
| BMI | 1.02 | .02 | .20 | .99 – 1.06 | 1.01 | .002 | <0.001 | 1.01 – 1.02 |
| QT Prolonging medication | 1.37 | 0.53 | 0.41 | 0.64 – 2.92 | 1.65 | 0.06 | <0.001 | 1.53 – 1.78 |
| Comorbid cardiovascular conditions | 2.13 | .30 | <0.001 | 1.61 – 2.81 | 1.45 | .03 | <0.001 | 1.41 – 1.49 |

HR = hazard ratio
SE = standard error
CI = confidence interval
QTc-PRS = polygenic risk score
SCD = Sudden Cardiac Death
BMI = body mass index
<sup>1</sup> HR per 5-unit change in continuous QTc-PRS
<sup>2</sup> HR per one year additional age
<sup>3</sup> HR for male compared to female
<sup>4</sup> Not available. No patient who died of SCD were on QT prolonging medication; no SCD deaths were observed among patients with sleep apnea who were Other race, Mixed race, or race not reported

The interaction term of QTc-PRS x diagnosed sleep apnea, which tests for effect modification by sleep apnea on the association between QTc-PRS and risk of death, was significant for SCD (p=0.01). While there was no association between QTc-PRS and SCD among participants without sleep apnea, the adjusted HR per 5 unit change in QTc-PRS for SCD among those with diagnosed sleep apnea was 1.64 (95% CI 1.16 - 2.31, p=0.005). We did not observe an association between QTc-PRS and all-cause death, with or without diagnosed sleep apnea (Table 3).

The mean (SD) QTc-PRS was markedly lower for White patients compared to other race categories: White 0.87 (7.4), Black 4.7 (5.7), Asian 5.3 (6.8), Other 3.4 (6.9), Mixed 2.5 (7.0), and Not Reported 2.1 (7.4). Among those with sleep apnea, Black patients (compared to White) had a significantly elevated risk of SCD (HR=9.6., 95% CI 1.24 - 74, p=0.03).

We observed an HR per 5-unit change in QTc-PRS of 1.67 (SE .33, 95% CI 1.14 – 2.44, p=0.009) for SCD among those with diagnosis of sleep apnea when race was an indicator for all non-White categories (including “not reported”) versus White (not shown in Table 3); the HR for non-White versus White was 5.2 (SE 4.1, 95% CI 1.09 – 24, p=0.04). Although there was no evidence of effect modification by race (White versus non-White) for SCD (p=0.86) or for SCD among those with sleep apnea (p =0.53), we repeated our analyses limited to White patients and found no appreciable difference in results. In White patients with diagnosed sleep apnea, the risk of SCD per 5-unit change in QTc-PRS was still increased with HR = 1.56 (SE .34, 95% CI 1.02 – 2.4, p=0.04). We again observed significant effect modification (interaction term) by diagnosed sleep apnea on the association between QTc-PRS and SCD when limited to White patients (p=.04).

The model for death due to SCD and QTc-PRS quintiles among only those with diagnosed sleep apnea could not converge properly. The reason is that there were no deaths from SCD in quintiles 1 and 2. Comparing quintiles 4/5 combined to 1/2/3 combined, the risk of SCD among those in quintiles 4/5 was HR=4.6 (SE=3.7, 95% CI 0.88 – 21, p=0.06).

Among those without sleep apnea, Asians had a significantly elevated risk of SCD associated with QTc-PRS compared to Whites (HR= 2.85, 95% CI 1.53 – 5.30, p=0.001). Being Black (HR= 0.80, 95% CI 0.69 – 0.93, p=0.004) or Asian (HR= 0.73, 95% CI .64-.83, p<0.001) seemed protective when compared to Whites for all-cause mortality associated with QTc-PRS (Table 3). The results of analyses performed with the additional causes of death are included in the supplementary data and yielded no additional findings of significance.

## DISCUSSION

QTc-PRS was closely associated with QTc duration in the UK Biobank. The QTc-PRS was independently associated with SCD with sleep apnea after adjustment for confounders (age, sex, BMI, race, QT prolonging medications and cardiovascular conditions). The risk of SCD was greatest among Black participants with diagnosed sleep apnea.

QTc-PRS was correlated with QTc duration in the UK Biobank, thus replicating the findings of QTGEN,^22^ QTSCD,^26^ Noseworthy et al.,^25^ and Arkin et al.^24^. While QTc duration and QTc-PRS seemed to have a linear relationship in participants without sleep apnea (n=24,057), we did not notice this linear relationship in those with sleep apnea likely due to the small number of patients with sleep apnea and QTc duration (n=213). As in Noseworthy et al. we also found that the mean QTc for the top QTc-PRS quintile is 15msec longer than the bottom quintile.

To our knowledge, Noseworthy et al., are the only other investigators who explored the relationship between QTc-PRS and SCD risk.^25^ Noseworthy et al. did not find a linear relationship between QTc-PRS and SCD risk in a Finnish population that lacked the diversity in race-ethnicity observed in our analysis of the UK Biobank. ^25^ In a secondary post-hoc analysis, Noseworthy et al. described a U-shaped relationship between QTc-PRS and SCD with risk of SCD higher in the top QTc-PRS quintile when compared to the middle quintile. ^25^ In our study no difference was noted in SCD risk between quintiles 1 and 2, increased SCD risk was noted for quintile 3 and again for quintile 4 with a plateau between quintiles 4 and 5. More research is needed to evaluate the correlation between QTc-PRS and SCD and to explore the influence of race and ethnicity on these relationships.

In our study, only four SNPs were associated with QTc duration when considered individually: rs12079745, rs3857067, rs1961102 and rs1052536 which mapped to *ATP1B1* (ATPase Na+/K+ transporting subunit beta 1), *HMGB3P15* (high mobility group box 3 pseudogene 15)/*ARAP2* (ArfGAP with RhoGAP domain, ankyrin repeat and PH domain 2), and *LIG 3* (DNA ligase 3) respectively. Other studies ^22,24–26^ have noted more SNPs to individually contribute to QTc which, which might correlate with the more heterogenous population of the UK Biobank population in regard to race and ethnicity when compared to other populations studied.

Noseworthy et al. described that none of the individual SNPs were associated with SCD risk in their study. In our study we found rs3934467 (NOS1AP) and rs3857067 (*HMGB3P15/ARAP2*) were individually correlated with SCD, although their p values reached only nominal significance. Neither of these two SNPs however were evaluated by Noseworthy et al. who at the time included 14 SNPs in the QTc-PRS while we have included additional SNPs since discovered to be associated with QTc in the QTc-PRS. ^24,27,35^

Most interestingly, sleep apnea was noted to be an effect modifier in the relationship between QTc-PRS and SCD. Patients with diagnosed sleep apnea and an elevated QTc-PRS were at increased risk for SCD. This intuitively makes sense given the propensity for patients with sleep apnea to independently manifest longer resting and sleep QTc duration, ^3,4^ which when combined with an increased genetic susceptibility of QT prolongation as measured by the QTc-PRS can lead to SCD. Black participants with sleep apnea were at the greatest risk for SCD perhaps due to increased prevalence and severity of sleep apnea in this population.^36–46^ Black people are also at a higher risk for QTc prolongation^47^ and SCD.^48^ These associations need to be validated in other studies.

The major limitation to this study is the small proportion of patients within the UK Biobank Cohort with QTc duration information which limits the power to detect associations with individual SNPs. Furthermore, there was a low number of participants in the death category of interest (SCD) which may have limited our ability to detect associations of the QTc-PRS with SCD. We also suspect that the prevalence of sleep apnea is underestimated given the global prevalence of sleep apnea^49^ even though the UK-Biobank cohort is thought to be healthier than the general population in the UK.^50^ This healthy selection bias likely limits the generalizability of the data to the general population.^50^ However, if our findings are significant in a healthy population, they may be even more pronounced in the sicker general population. We also were unable to differentiate between obstructive and central sleep apnea, although, it can be assumed that a majority of the patients had obstructive sleep apnea since less than one percent of the population are thought to have central sleep apnea.^33^ The hypoxic burden of sleep apnea and severity of apnea is also not known for the sleep apnea cohort given the absence of sleep study data in the database. Severity of obstructive sleep apnea and hypoxic burden has been associated with cardiovascular disease.^51^ The advantage of this study is the use of a large population-based cohort with readily available mortality data from national death registries, along with linked genetic data.

## CONCLUSIONS

Our study demonstrates an association between QTc-PRS and QTc duration in the UK Biobank population. The QTc-PRS was associated with SCD, but only among participants with sleep apnea, and Black participants with sleep apnea had a particularly high risk of SCD.

## AUTHORS AND CONTRIBUTORS

AA: the acquisition, analysis, and interpretation of data; revising manuscript, final approval of the version to be published; agreement to be accountable for all aspects of the work.

SIP: conception and design of the work; the acquisition, analysis, and interpretation of data; drafting and revising manuscript, final approval of the version to be published; agreement to be accountable for all aspects of the work.

WZ, RLW, YCK, IYP, SFQ, FS, SG, SP: conception and design of the work; interpretation of data; revising manuscript, final approval of the version to be published; agreement to be accountable for all aspects of the work.

CW: analysis, and interpretation of data; revising manuscript, final approval of the version to be published; agreement to be accountable for all aspects of the work.

## ABBREVIATIONS

QTc: Corrected QT interval
QTc-PRS: polygenic risk score for QT prolongation
SCD: Sudden cardiac death

## Supplementary Appendix

## Data Availability

All data can be made available as needed in accordance with any limitations as per the UK Biobank data share agreement.

https://biobank.ndph.ox.ac.uk/

---

**Table S1:**
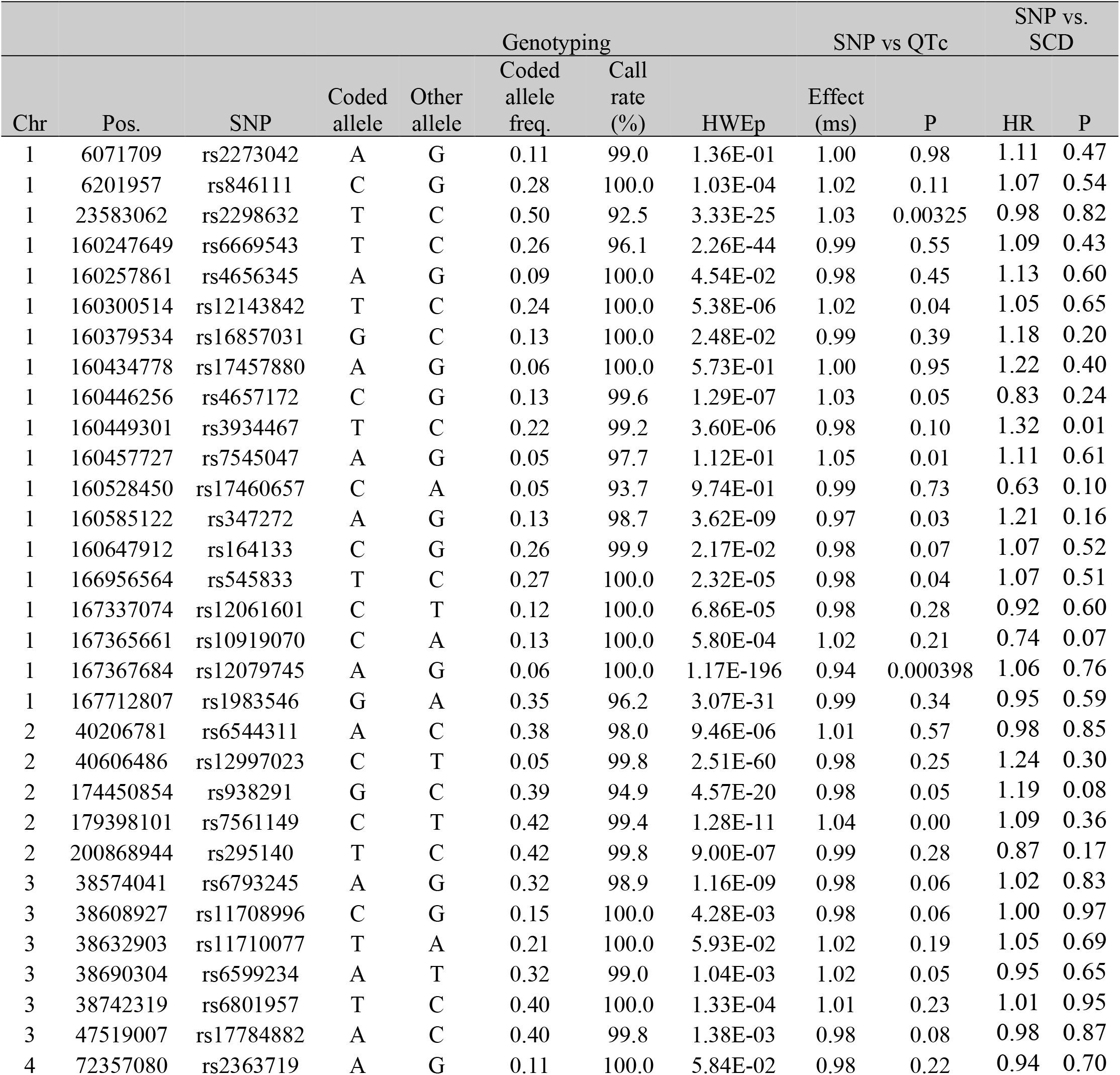
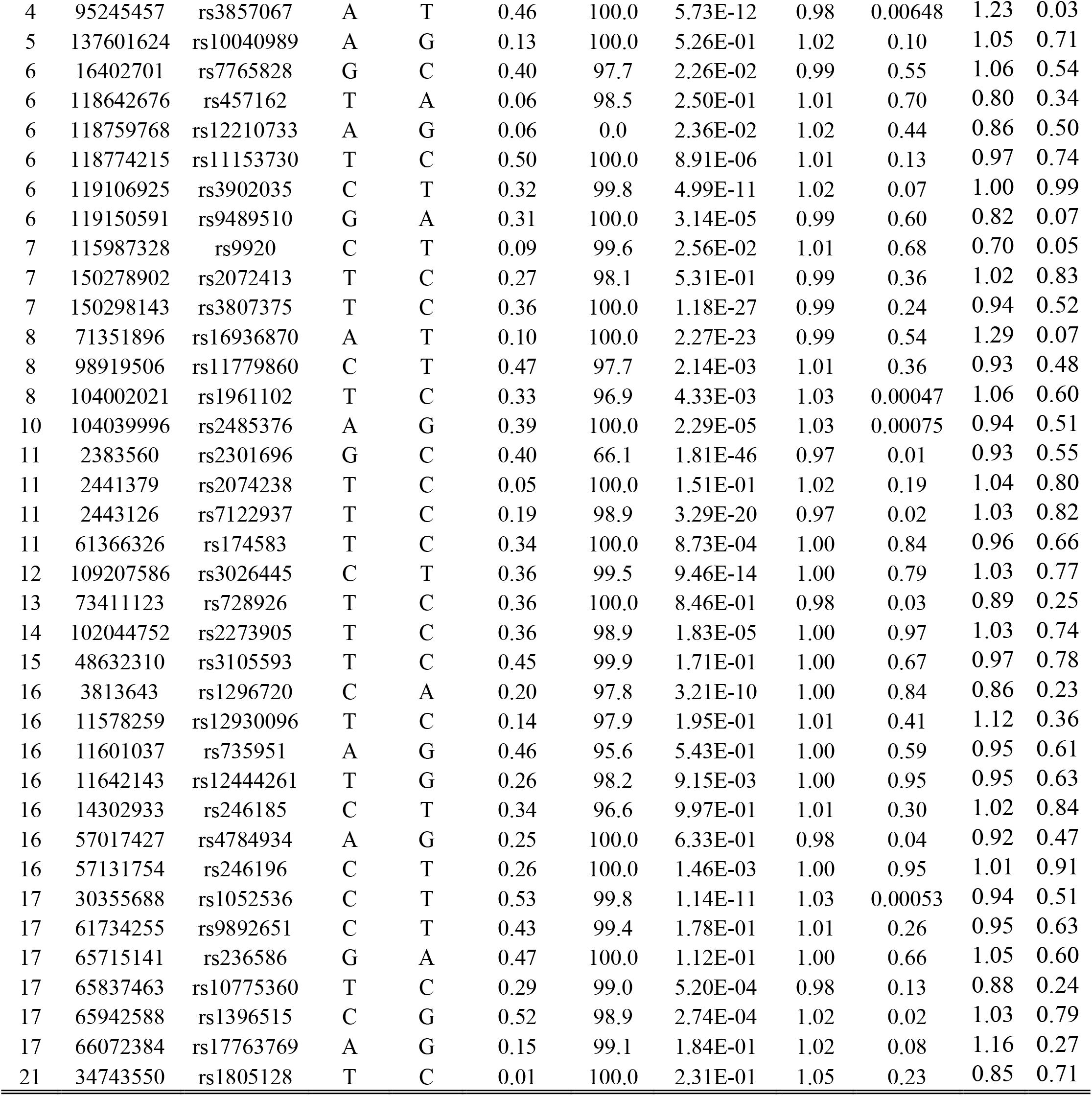
Individual SNP effects for changes in QTc and Sudden Cardiac Death (SCD)

**Table S2.** Participant demographics with additional mortality categories in the UK Biobank.

| Characteristic | SCD<sup>1</sup><br>(n= 223) | Probable<br>SCD<sup>2</sup><br>(n= 155) | Possible<br>SCD<sup>3</sup><br>(n= 6304) | Other Death<sup>4</sup><br>(n= 12,926) | Death from<br>all causes<br>(n= 19,589) | Alive<br>(n= 467,691) |
| --- | --- | --- | --- | --- | --- | --- |
| Age, mean (SD)<br>[range] | 60.8 (6.9)<br>[42 to 70] | 62.3 (6.3)<br>[40 to 70] | 62.2 (6.1)<br>[42 to 70] | 61.1 (6.7)<br>[40 to 70] | 61.4 (6.5)<br>[40 to 70] | 56.3 (8.1)<br>[37 to 73] |
| Age > 50 years,<br>count (%) | 196 (87.9) | 143 (92.3) | 5,897 (93.5) | 11,741 (90.8) | 17,961 (91.7) | 341,392 (73.0) |
| Sex, count (%) |  |  |  |  |  |  |
| Male | 154 (69.1) | 100 (64.5) | 4,468 (70.9) | 7,149 (55.3) | 11,856 (60.5) | 211,217 (45.2) |
| Female | 69 (31.1) | 55 (35.5) | 1,836 (29.1) | 5,777 (44.7) | 7,733 (39.5) | 256,474 (54.8) |
| Race, count (%) |  |  |  |  |  |  |
| White | 198 (88.8) | 147 (94.8) | 6,007 (95.3) | 12473 (96.5) | 18806 (96.0) | 440,405 (94.2) |
| Black | 4 (1.8) | 0 (0.0) | 66 (1.1) | 111 (0.9) | 181 (0.9) | 7,463 (1.6) |
| Asian | 15 (6.7) | 2 (1.3) | 131 (2.1) | 126 (1.0) | 274 (1.4) | 10,646 (2.3) |
| Other | 3 (1.4) | 3 (1.9) | 33 (0.5) | 80 (0.6) | 119 (0.6) | 4,235 (0.9) |
| Mixed | 1 (0.5) | 1 (0.7) | 26 (0.4) | 48 (0.4) | 76 (0.4) | 2,767 (0.6) |
| Not Reported | 2 (0.9) | 2 (1.3) | 41 (0.7) | 88 (0.7) | 133 (0.7) | 2,175 (0.5) |
| BMI, mean (SD) | 28.8 (6.0) | 28.7 (5.6) | 29.2 (5.8) | 27.7 (5.1) | 28.2 (5.4) | 27.4 (4.8) |
| QTc-PRS<sup>5</sup>, mean<br>(SD) | 1.9 (7.3) | .67 (7.8) | .91 (7.4) | 1.1 (7.5) | 1.0 (7.4) | 1.1 (7.4) |
| QTc-PRS<sup>5</sup> quintile<br>cut-offs |  |  |  |  |  |  |
| Minimum | -19.2 | -17.6 | -24.9 | -28.6 | -28.6 | -33.9 |
| 20 percentile | - 4.4 | - 6.4 | - 5.3 | - 5.1 | - 5.2 | - 5.2 |
| 40 percentile | -0.37 | -1.5 | -0.94 | -0.91 | -0.91 | -0.86 |
| 60 percentile | 3.87 | 2.1 | 2.7 | 2.83 | 2.8 | 2.9 |
| 80 percentile | 7.8 | 7.7 | 7.1 | 7.24 | 7.2 | 7.3 |
| Maximum | 22.8 | 18.2 | 35.6 | 28.2 | 35.6 | 36.8 |
| Sleep apnea<br>diagnosed, count<br>(%) | 8 (3.6) | 8 (5.2) | 248 (3.9) | 265 (2.1) | 529 (2.7) | 6,117 (1.3) |
| Sleep apnea self-reported, count (%) | 1 (0.45) | 1 (0.65) | 49 (0.78) | 47 (.36) | 98 (0.50) | 1,483 (0.32) |
| Prescribed QT<br>Prolonging<br>medication (%) | 7 (3.1) | 14 (9.0) | 275 (4.4) | 496 (3.8) | 792 (4.0) | 12,213 (2.6) |
| Comorbid<br>cardiovascular<br>conditions, count<br>(%) | 126 (56.8) | 80 (52.0) | 4,002 (63.7) | 5,065 (39.4) | 9,267 (47.5) | 135,445 (29.1) |

SD = standard deviation
<sup>1</sup>SCD = Sudden Cardiac Death
<sup>2</sup>Probable SCD = all other conduction disorders excluding SCD
<sup>3</sup> Possible SCD = all other cardiovascular causes not including SCD and probable SCD
<sup>4</sup> Other death = deaths from non-cardiovascular causes
<sup>5</sup> QTc-PRS = QTc polygenic risk score

**Table S3.** Risk of death associated with QTc-PRS in additional mortality categories.

| Cause of death | Unadjusted |  |  |  | Adjusted<sup>2</sup> |  |  |  |
| --- | --- | --- | --- | --- | --- | --- | --- | --- |
|  | HR<sup>1</sup> | SE | p-value | 95% CI | HR<sup>1</sup> | SE | p-value | 95% CI |
| SCD | 1.08 | 0.05 | 0.10 | 0.99 - 1.17 | 1.05 | 0.05 | 0.29 | 0.96 - 1.15 |
| Probable SCD | 0.96 | 0.05 | 0.52 | 0.86 - 1.08 | 0.97 | 0.06 | 0.63 | 0.87 - 1.09 |
| Possible SCD | 0.99 | 0.008 | 0.09 | 0.97 – 1.00 | 0.99 | 0.009 | 0.22 | 0.97 – 1.01 |
| Other death | 1.00 | 0.006 | 0.90 | 0.99–1.01 | 1.01 | 0.006 | 0.20 | 1.00–1.02 |
| All causes | 1.00 | 0.005 | 0.31 | 0.99–1.00 | 1.00 | 0.005 | 0.70 | 0.99–1.01 |

HR = hazard ratio
SE = standard error
CI = confidence interval
QTc-PRS = QTc polygenic risk score
SCD = Sudden Cardiac Death
Probable SCD = all conduction disorders excluding SCD
Possible SCD = all other cardiovascular causes not including SCD and probable SCD
Other death = deaths from non-cardiovascular causes
<sup>1</sup> HR per 5-unit change in continuous QTc-PRS
<sup>2</sup> Adjusted for age, sex, race, BMI, QT Prolonging medication, and comorbid cardiovascular conditions

**Table S4:**
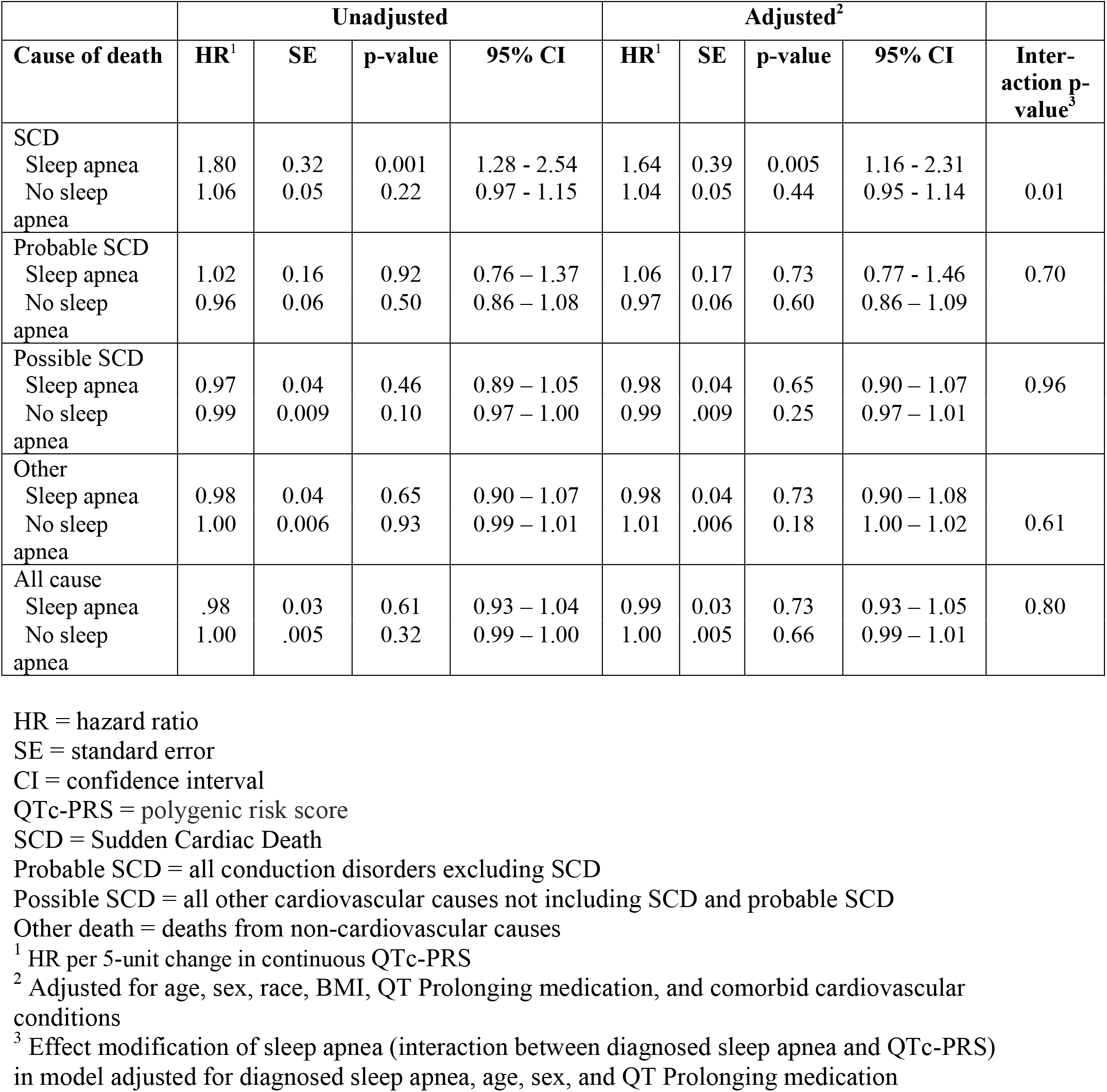
Risk of all death categories associated with QTc-PRS, stratified by sleep apnea.

## REFERENCES

1. Patel SI, Zareba W, Wendel C, et al. A QTc risk score in patients with obstructive sleep apnea. Sleep Med 2023;103:159–64.

2. Shamsuzzaman A, Amin RS, van der Walt C, et al. Daytime cardiac repolarization in patients with obstructive sleep apnea. Sleep Breath 2015;19:1135–40.

3. Sokmen E, Ozbek SC, Celik M, Sivri S, Metin M, Avcu M. Changes in the parameters of ventricular repolarization during preapnea, apnea, and postapnea periods in patients with obstructive sleep apnea. Pacing Clin Electrophysiol 2018.

4. Gillis AM, Stoohs R, Guilleminault C. Changes in the QT interval during obstructive sleep apnea. Sleep 1991;14:346–50.

5. Patel SI, Zareba W, LaFleur B, et al. Markers of ventricular repolarization and overall mortality in sleep disordered breathing. Sleep Med 2022;95:9–15.

6. Moss AJ. The QT interval and torsade de pointes. Drug Saf 1999;21 Suppl 1:5–10; discussion 81-7.

7. Tisdale JE, Wroblewski HA, Overholser BR, Kingery JR, Trujillo TN, Kovacs RJ. Prevalence of QT interval prolongation in patients admitted to cardiac care units and frequency of subsequent administration of QT interval-prolonging drugs: a prospective, observational study in a large urban academic medical center in the US. Drug Saf 2012;35:459–70.

8. Straus SM, Kors JA, De Bruin ML, et al. Prolonged QTc interval and risk of sudden cardiac death in a population of older adults. J Am Coll Cardiol 2006;47:362–7.

9. Hobbs JB, Peterson DR, Moss AJ, et al. Risk of aborted cardiac arrest or sudden cardiac death during adolescence in the long-QT syndrome. Jama 2006;296:1249–54.

10. Zhang Y, Post WS, Blasco-Colmenares E, Dalal D, Tomaselli GF, Guallar E. Electrocardiographic QT interval and mortality: a meta-analysis. Epidemiology 2011;22:660–70.

11. Sohaib SM, Papacosta O, Morris RW, Macfarlane PW, Whincup PH. Length of the QT interval: determinants and prognostic implications in a population-based prospective study of older men. J Electrocardiol 2008;41:704–10.

12. Freeman BD, Dixon DJ, Coopersmith CM, Zehnbauer BA, Buchman TG. Pharmacoepidemiology of QT-interval prolonging drug administration in critically ill patients. Pharmacoepidemiol Drug Saf 2008;17:971–81.

13. Chao CC, Wang TL, Chong CF, et al. Prognostic value of QT parameters in patients with acute hemorrhagic stroke: a prospective evaluation with respect to mortality and post-hospitalization bed confinement. J Chin Med Assoc 2009;72:124–32.

14. Dekker JM, Crow RS, Hannan PJ, Schouten EG, Folsom AR. Heart rate-corrected QT interval prolongation predicts risk of coronary heart disease in black and white middle-aged men and women: the ARIC study. J Am Coll Cardiol 2004;43:565–71.

15. Okin PM, Devereux RB, Howard BV, Fabsitz RR, Lee ET, Welty TK. Assessment of QT interval and QT dispersion for prediction of all-cause and cardiovascular mortality in American Indians: The Strong Heart Study. Circulation 2000;101:61–6.

16. Salles GF, Cardoso CR, Muxfeldt ES. Prognostic value of ventricular repolarization prolongation in resistant hypertension: a prospective cohort study. J Hypertens 2009;27:1094–101.

17. Robbins J, Nelson JC, Rautaharju PM, Gottdiener JS. The association between the length of the QT interval and mortality in the Cardiovascular Health Study. Am J Med 2003;115:689–94.

18. Montanez A, Ruskin JN, Hebert PR, Lamas GA, Hennekens CH. Prolonged QTc interval and risks of total and cardiovascular mortality and sudden death in the general population: a review and qualitative overview of the prospective cohort studies. Arch Intern Med 2004;164:943–8.

19. Pickham D, Helfenbein E, Shinn JA, et al. High prevalence of corrected QT interval prolongation in acutely ill patients is associated with mortality: results of the QT in Practice (QTIP) Study. Crit Care Med 2012;40:394–9.

20. Patel SI, Ackerman MJ, Shamoun FE, et al. QT prolongation and sudden cardiac death risk in hypertrophic cardiomyopathy. Acta Cardiol 2019;74:53–8.

21. Akylbekova EL, Crow RS, Johnson WD, et al. Clinical correlates and heritability of QT interval duration in blacks: the Jackson Heart Study. Circ Arrhythm Electrophysiol 2009;2:427–32.

22. Newton-Cheh C, Eijgelsheim M, Rice KM, et al. Common variants at ten loci influence QT interval duration in the QTGEN Study. Nat Genet 2009;41:399–406.

23. Hong Y, Rautaharju PM, Hopkins PN, et al. Familial aggregation of QT-interval variability in a general population: results from the NHLBI Family Heart Study. Clin Genet 2001;59:171–7.

24. Arking DE, Pulit SL, Crotti L, et al. Genetic association study of QT interval highlights role for calcium signaling pathways in myocardial repolarization. Nat Genet 2014;46:826–36.

25. Noseworthy PA, Havulinna AS, Porthan K, et al. Common genetic variants, QT interval, and sudden cardiac death in a Finnish population-based study. Circ Cardiovasc Genet 2011;4:305–11.

26. Pfeufer A, Sanna S, Arking DE, et al. Common variants at ten loci modulate the QT interval duration in the QTSCD Study. Nat Genet 2009;41:407–14.

27. Strauss DG, Vicente J, Johannesen L, et al. Common Genetic Variant Risk Score Is Associated With Drug-Induced QT Prolongation and Torsade de Pointes Risk: A Pilot Study. Circulation 2017;135:1300–10.

28. About UK Biobank. (Accessed September 1, 2019, at https://www.ukbiobank.ac.uk/about-biobank-uk/)

29. UK Biobank Axiom® Array Content Summary. (Accessed September 1, 2019, at http://www.ukbiobank.ac.uk/wp-content/uploads/2014/04/uk-biobank-axiom-array-content-summary-2014-1.pdf.)

30. Haugaa KH, Bos JM, Tarrell RF, Morlan BW, Caraballo PJ, Ackerman MJ. Institution-wide QT alert system identifies patients with a high risk of mortality. Mayo Clin Proc 2013;88:315–25.

31. Li Y, Willer C, Sanna S, Abecasis G. Genotype imputation. Annu Rev Genomics Hum Genet 2009;10:387–406.

32. Cuzick J. A Wilcoxon-type test for trend. Statistics in Medicine 1985;4:87–90.

33. Donovan LM, Kapur VK. Prevalence and Characteristics of Central Compared to Obstructive Sleep Apnea: Analyses from the Sleep Heart Health Study Cohort. Sleep 2016;39:1353–9.

34. Fine JP, Gray RJ. A proportional hazards model for the subdistribution of a competing risk. J Am Stat Assoc 1999;94:496–509.

35. Turkowski KL, Dotzler SM, Tester DJ, et al. Corrected QT Interval-Polygenic Risk Score and Its Contribution to Type 1, Type 2, and Type 3 Long-QT Syndrome in Probands and Genotype-Positive Family Members. Circ Genom Precis Med 2020;13:e002922.

36. Alkhazna A, Bhat A, Ladesich J, Barthel B, Bohnam AJ. Severity of obstructive sleep apnea between black and white patients. Hosp Pract (1995) 2011;39:82–6.

37. Ancoli-Israel S, Klauber MR, Stepnowsky C, Estline E, Chinn A, Fell R. Sleep-disordered breathing in African-American elderly. Am J Respir Crit Care Med 1995;152:1946–9.

38. Kripke DF, Ancoli-Israel S, Klauber MR, Wingard DL, Mason WJ, Mullaney DJ. Prevalence of sleep-disordered breathing in ages 40-64 years: a population-based survey. Sleep 1997;20:65–76.

39. Nieto FJ, Young TB, Lind BK, et al. Association of sleep-disordered breathing, sleep apnea, and hypertension in a large community-based study. Sleep Heart Health Study. JAMA 2000;283:1829–36.

40. Redline S, Tishler PV, Hans MG, Tosteson TD, Strohl KP, Spry K. Racial differences in sleep-disordered breathing in African-Americans and Caucasians. Am J Respir Crit Care Med 1997;155:186–92.

41. Redline S, Tishler PV, Schluchter M, Aylor J, Clark K, Graham G. Risk factors for sleep-disordered breathing in children. Associations with obesity, race, and respiratory problems. Am J Respir Crit Care Med 1999;159:1527–32.

42. Rosen CL, Larkin EK, Kirchner HL, et al. Prevalence and risk factors for sleep-disordered breathing in 8- to 11-year-old children: association with race and prematurity. J Pediatr 2003;142:383–9.

43. Ruiter ME, DeCoster J, Jacobs L, Lichstein KL. Sleep disorders in African Americans and Caucasian Americans: a meta-analysis. Behav Sleep Med 2010;8:246–59.

44. Stepanski E, Zayyad A, Nigro C, Lopata M, Basner R. Sleep-disordered breathing in a predominantly African-American pediatric population. J Sleep Res 1999;8:65–70.

45. Weinstock TG, Rosen CL, Marcus CL, et al. Predictors of obstructive sleep apnea severity in adenotonsillectomy candidates. Sleep 2014;37:261–9.

46. Young T, Shahar E, Nieto FJ, et al. Predictors of sleep-disordered breathing in community-dwelling adults: the Sleep Heart Health Study. Arch Intern Med 2002;162:893–900.

47. Williams ES, Thomas KL, Broderick S, et al. Race and gender variation in the QT interval and its association with mortality in patients with coronary artery disease: results from the Duke Databank for Cardiovascular Disease (DDCD). Am Heart J 2012;164:434–41.

48. Zhao D, Post WS, Blasco-Colmenares E, et al. Racial Differences in Sudden Cardiac Death. Circulation 2019;139:1688–97.

49. Benjafield AV, Ayas NT, Eastwood PR, et al. Estimation of the global prevalence and burden of obstructive sleep apnoea: a literature-based analysis. Lancet Respir Med 2019;7:687–98.

50. Fry A, Littlejohns TJ, Sudlow C, et al. Comparison of Sociodemographic and Health-Related Characteristics of UK Biobank Participants With Those of the General Population. Am J Epidemiol 2017;186:1026–34.

51. Azarbarzin A, Sands SA, Stone KL, et al. The hypoxic burden of sleep apnoea predicts cardiovascular disease-related mortality: the Osteoporotic Fractures in Men Study and the Sleep Heart Health Study. Eur Heart J 2019;40:1149–57.

